# Tocilizumab in Hospitalized Patients With COVID-19 Pneumonia

**DOI:** 10.1101/2020.08.27.20183442

**Authors:** Ivan O. Rosas, Norbert Bräu, Michael Waters, Ronaldo Go, Bradley D. Hunter, Sanjay Bhagani, Daniel Skiest, Mariam S. Aziz, Nichola Cooper, Ivor S. Douglas, Sinisa Savic, Taryn Youngstein, Lorenzo Del Sorbo, Antonio Cubillo Gracian, David J. De La Zerda, Andrew Ustianowski, Min Bao, Sophie Dimonaco, Emily Graham, Balpreet Matharu, Helen Spotswood, Larry Tsai, Atul Malhotra

**Author notes:** **Corresponding author:** Ivan O. Rosas, M.D. Professor and Section Chief, Pulmonary, Critical Care, and Sleep Medicine, Baylor College of Medicine, 7200 Cambridge Street, Houston, Texas 77030.

## Abstract

**BACKGROUND:** COVID-19 is associated with immune dysregulation and hyperinflammation. Tocilizumab is an anti–interleukin-6 receptor antibody.

**METHODS:** Patients hospitalized with severe COVID-19 pneumonia receiving standard care were randomized (2:1) to double-blinded intravenous tocilizumab 8 mg/kg or placebo. The primary outcome measure was clinical status on a 7-category ordinal scale at day 28 (1, discharged/ready for discharge; 7, death).

**RESULTS:** Overall, 452 patients were randomized; the modified-intention-to-treat population included 294 tocilizumab-treated and 144 placebo-treated patients. Clinical status at day 28 was not statistically significantly improved for tocilizumab versus placebo (P=0.36). Median (95% CI) ordinal scale values at day 28: 1.0 (1.0 to 1.0) for tocilizumab and 2.0 (1.0 to 4.0) for placebo (odds ratio, 1.19 [0.81 to 1.76]). There was no difference in mortality at day 28 between tocilizumab (19.7%) and placebo (19.4%) (difference, 0.3% [95% CI, –7.6 to 8.2]; nominal P=0.94). Median time to hospital discharge was 8 days shorter with tocilizumab than placebo (20.0 and 28.0, respectively; nominal P=0.037; hazard ratio 1.35 [95% CI 1.02 to 1.79]). Median duration of ICU stay was 5.8 days shorter with tocilizumab than placebo (9.8 and 15.5, respectively; nominal P=0.045). In the safety population, serious adverse events occurred in 34.9% of 295 patients in the tocilizumab arm and 38.5% of 143 in the placebo arm.

**CONCLUSIONS:** In this randomized placebo-controlled trial in hospitalized COVID-19 pneumonia patients, tocilizumab did not improve clinical status or mortality. Potential benefits in time to hospital discharge and duration of ICU stay are being investigated in ongoing clinical trials.

**Trial registration:** ClinicalTrials.gov NCTG432G615

---

Coronavirus disease 2019 (COVID-19) has rapidly developed into a global health threat since emerging in China in late 2019.^1^ Severe COVID-19 pneumonia, occurring in approximately 15% of patients infected with severe acute respiratory syndrome coronavirus-2 (SARS-CoV-2), is associated with high mortality rates and places extensive burden on intensive care units to provide mechanical ventilation and other advanced forms of life support.^2,3^ Similar to Middle Eastern Respiratory Syndrome and SARS-CoV-1,^4^ an initial phase of COVID-19 with high viral replication precedes a second disease phase that may be driven by the host immune response. This can lead to rapid increase in proinflammatory cytokines, an uncontrolled inflammatory response, acute respiratory distress syndrome (ARDS), and multiple organ failure.^4,5^ Interleukin-6 levels correlate with COVID-19 severity,^6,7^ suggesting that, in this setting, immune dysregulation and ARDS might be influenced by interleukin-6.^5,8^ Accumulation of lymphocytes and inflammatory monocytes, endotheliitis, apoptosis, thrombosis, and angiogenesis in the pulmonary vasculature of patients with COVID-19 suggests that vascular inflammation and dysfunction contribute to the pathophysiology of severe COVID-19 pneumonia.^9,10^ Interleukin-6 promotes endothelial dysfunction and development of vascular permeability and might play a role in the vascular dysfunction of this disease.^11^

The potential role of interleukin-6 in COVID-19 pneumonia^5,8^ provides rationale for investigation of interleukin-6 signaling inhibitors. Tocilizumab is a monoclonal anti–interleukin-6 receptor-alpha blocking antibody used to treat certain inflammatory diseases.^12^ Improvements in patients with severe COVID-19 pneumonia who received tocilizumab were observed in case reports^13-15^ and supported by retrospective observational cohort studies that showed rapid reduction in fever, reduced need for oxygen support and mechanical ventilation, and improvement in lung manifestations.^16-21^

This is the first global, randomized, double-blind, placebo-controlled trial to investigate whether tocilizumab has clinical benefit in hospitalized patients with severe COVID-19 pneumonia.

## PATIENTS AND METHODS

### Trial Design and Oversight

COVACTA is a global, multicenter, randomized, double-blind, placebo-controlled, phase 3 trial investigating the efficacy and safety of tocilizumab in patients with severe COVID-19 pneumonia (ClinicalTrials.gov, NCT04320615). Patients 18 years or older with severe COVID-19 pneumonia confirmed by positive polymerase chain reaction test in any body fluid and evidenced by bilateral chest infiltrates on chest x-ray or computed tomography were enrolled. Eligible patients had blood oxygen saturation ≤93% or partial pressure of oxygen/fraction of inspired oxygen <300 mm/Hg. Patients were excluded if the treating physician determined that death was imminent and inevitable within 24 hours or if they had active tuberculosis or bacterial, fungal, or viral infection other than SARS-CoV-2. Standard care per local practice (antiviral treatment, low-dose steroids, convalescent plasma, supportive care) was permitted; however, concomitant treatment with another investigational agent (except antivirals) or any immunomodulatory agent was prohibited. Informed consent was obtained for all enrolled patients. The study was conducted in accordance with the International Council for Harmonization E6 guideline for good clinical practice and the Declaration of Helsinki or local regulations, whichever afforded greater patient protection. The protocol was reviewed by institutional review boards or ethics committees.

Eligible patients were randomized (2:1) to receive intravenous tocilizumab (8 mg/kg infusion, maximum 800 mg) or placebo plus standard care using an interactive voice or web-based response system and permuted-block randomization. Randomization was stratified by geographic region (North America, Europe) and mechanical ventilation (yes, no). If clinical signs or symptoms did not improve or worsened (defined as sustained fever or worsened ordinal scale clinical status), a second infusion could be administered 8 to 24 hours after the first. The primary analysis was performed at day 28, and the final study visit occurred at day 60.

### Outcome Measures

The primary efficacy outcome was clinical status assessed on a 7-category ordinal scale (1, discharged or ready for discharge; 2, non–intensive care unit [ICU] hospital ward, not requiring supplemental oxygen; 3, non–ICU hospital ward, requiring supplemental oxygen; 4, ICU or non–ICU hospital ward, requiring noninvasive ventilation or high-flow oxygen; 5, ICU, requiring intubation and mechanical ventilation; 6, ICU, requiring extracorporeal membrane oxygenation or mechanical ventilation and additional organ support; 7, death) at day 28. Clinical status was recorded at baseline and every day during hospitalization. Key secondary efficacy endpoints were clinical status at day 14 on the 7-category ordinal scale, mortality at day 28, ventilator-free days to day 28, time to improvement from baseline in ≥2 categories on the 7-category ordinal scale, and time to hospital discharge (or ready for discharge [defined as normal body temperature and respiratory rate and stable oxygen saturation on ambient air or ≤2 L supplemental oxygen]). Other secondary endpoints included time to clinical failure defined as death, withdrawal during hospitalization, mechanical ventilation, or ICU transfer (for patients intubated or in the ICU at baseline, a 1-category worsening of clinical status was considered clinical failure); incidence of mechanical ventilation (among those not mechanically ventilated at randomization); incidence of ICU transfer (among those not in ICUs at baseline); and duration of ICU stay. Adverse events were recorded according to Medical Dictionary for Regulatory Activities system organ class and preferred term.

### Statistical Analysis

Efficacy was assessed in the modified-intention-to-treat (mITT) population (any randomized patients who received study medication) for the primary and secondary endpoints according to treatment assigned at randomization. Analyses were stratified by region and mechanical ventilation status at randomization except for some subgroup analyses, as specified. The primary endpoint compared distribution of the ordinal scale of clinical status between treatment groups using a nonparametric van Elteren test. The ratio of the odds of being in a better clinical status category for tocilizumab versus placebo was determined using a proportional odds model to give odds ratios and 95% CIs. Data from the last available postbaseline assessment on the ordinal scale were used for patients who withdrew before day 28, and all deaths and hospital discharges were carried forward. Differences in mortality were analyzed using the Cochran-Mantel-Haenszel test, differences in the number of ventilator-free days were assessed using the van Elteren test, and time-to-event secondary endpoints were assessed using a log-rank test with Kaplan-Meier plots produced (deaths were right-censored for all time-to-event endpoints assessing improvement). Cumulative incidence plots were generated using the nonparametric Aalen-Johansen estimator, where death is a competing risk. Safety was assessed in the safety-evaluable population (all patients who received any study medication) according to treatment first received. An estimated mITT population sample size of 450 patients randomized to tocilizumab or placebo was determined to give 90% power for the primary endpoint using the van Elteren test and an assumed distribution of the ordinal scale (Appendix 2). If significance was met, mortality at day 28 would be tested at the 5% level, but no other adjustment for multiplicity was planned.

## RESULTS

### Patients

Overall, 479 patients from 9 countries (Canada, Denmark, France, Germany, Italy, Netherlands, Spain, United Kingdom, United States) were screened, 452 patients were randomized, and 438 received study treatment (Figure 1). The mITT population included 294 patients randomized to tocilizumab and 144 to placebo. The safety population included 295 and 143 patients, respectively, because 1 patient randomized to placebo received tocilizumab. Overall, 224 of 301 patients (74.4%) randomized to tocilizumab and 108 of 151 patients (71.5%) randomized to placebo completed the 28-day follow-up. Excluding those who died, 20 patients (6.6%) from the tocilizumab arm and 14 (9.3%) from the placebo arm discontinued before day 28; none discontinued because of safety reasons.

**Figure 1.**
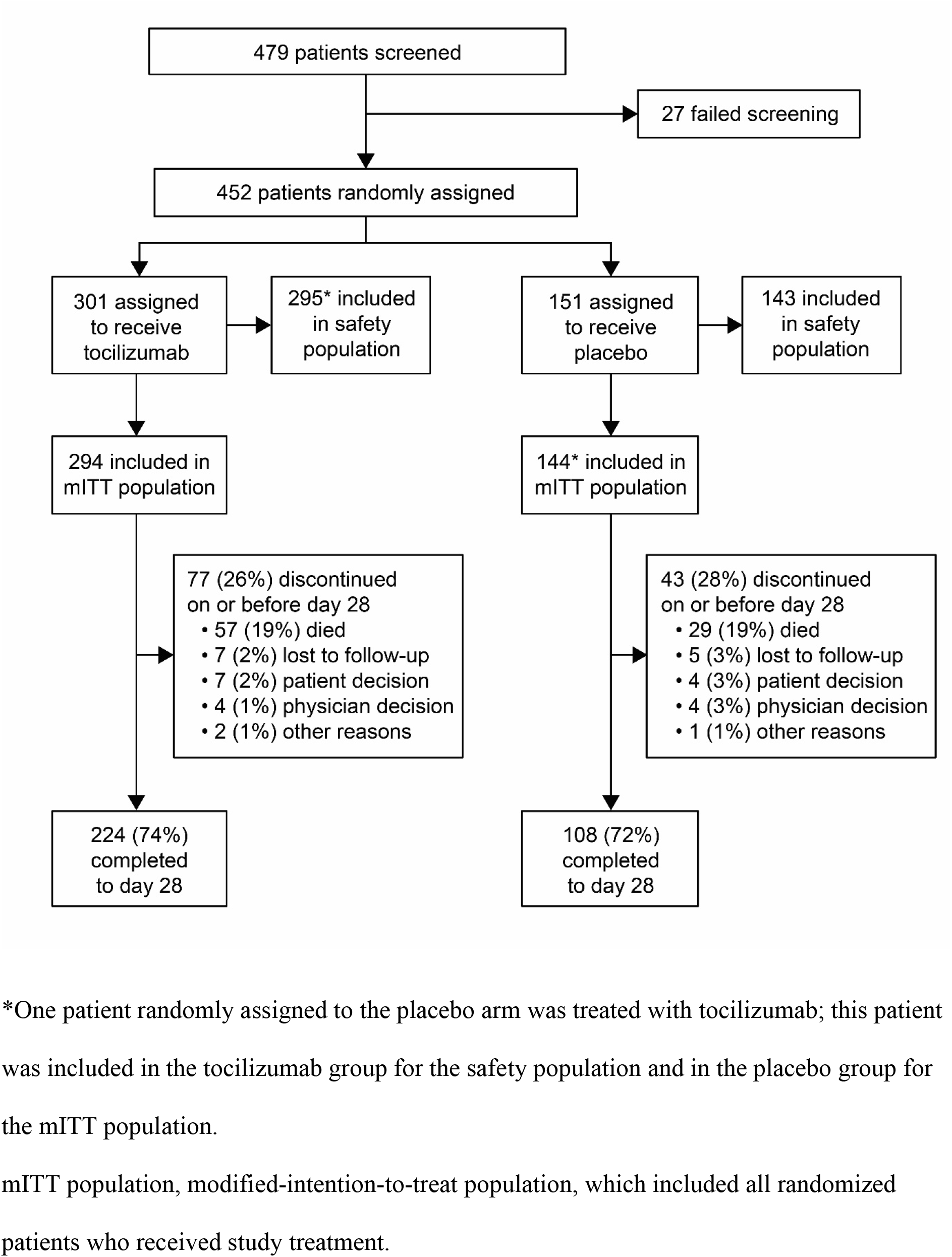
Patient disposition.

Baseline demographics and disease characteristics were generally balanced between treatment arms. Approximately 70% of patients in each arm were men; 176 patients (59.9%) were white and 40 (13.6%) were black in the tocilizumab arm compared with 76 (52.8%) and 26 (18.1%), respectively, in the placebo arm. Mean age was 60.9 ±14.6 years in the tocilizumab arm and 60.6 ±13.7 years in the placebo arm.

At baseline or any time during the study, lower proportions of patients in the tocilizumab than the placebo arm received steroids (106 [36.1%] vs 79 [54.9%]), antiviral treatment (87 [29.6%] vs 51 [35.4%]), and convalescent plasma (10 [3.4%] vs 6 [4.2%]) (Table 1).

**Table 1.**
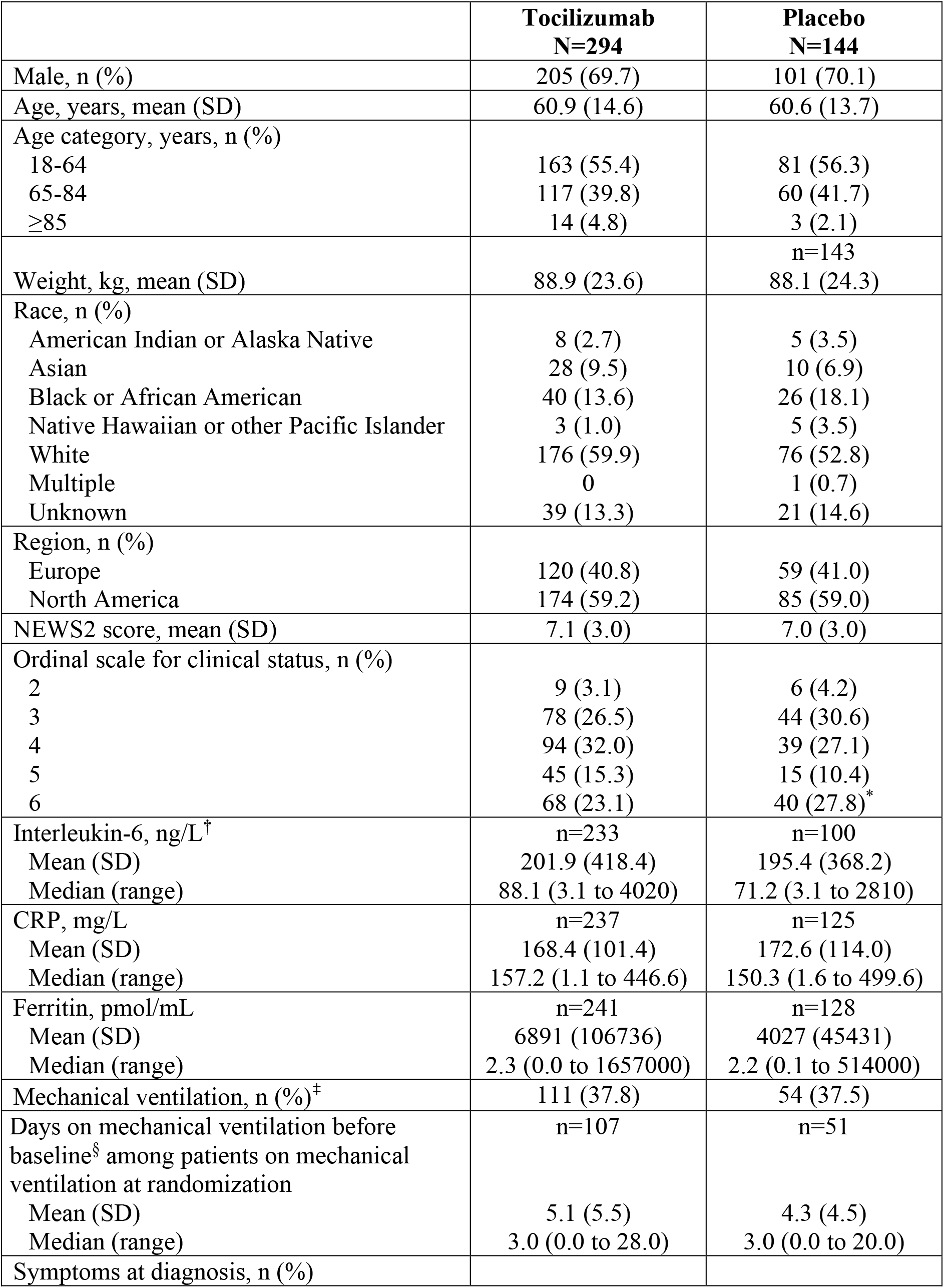
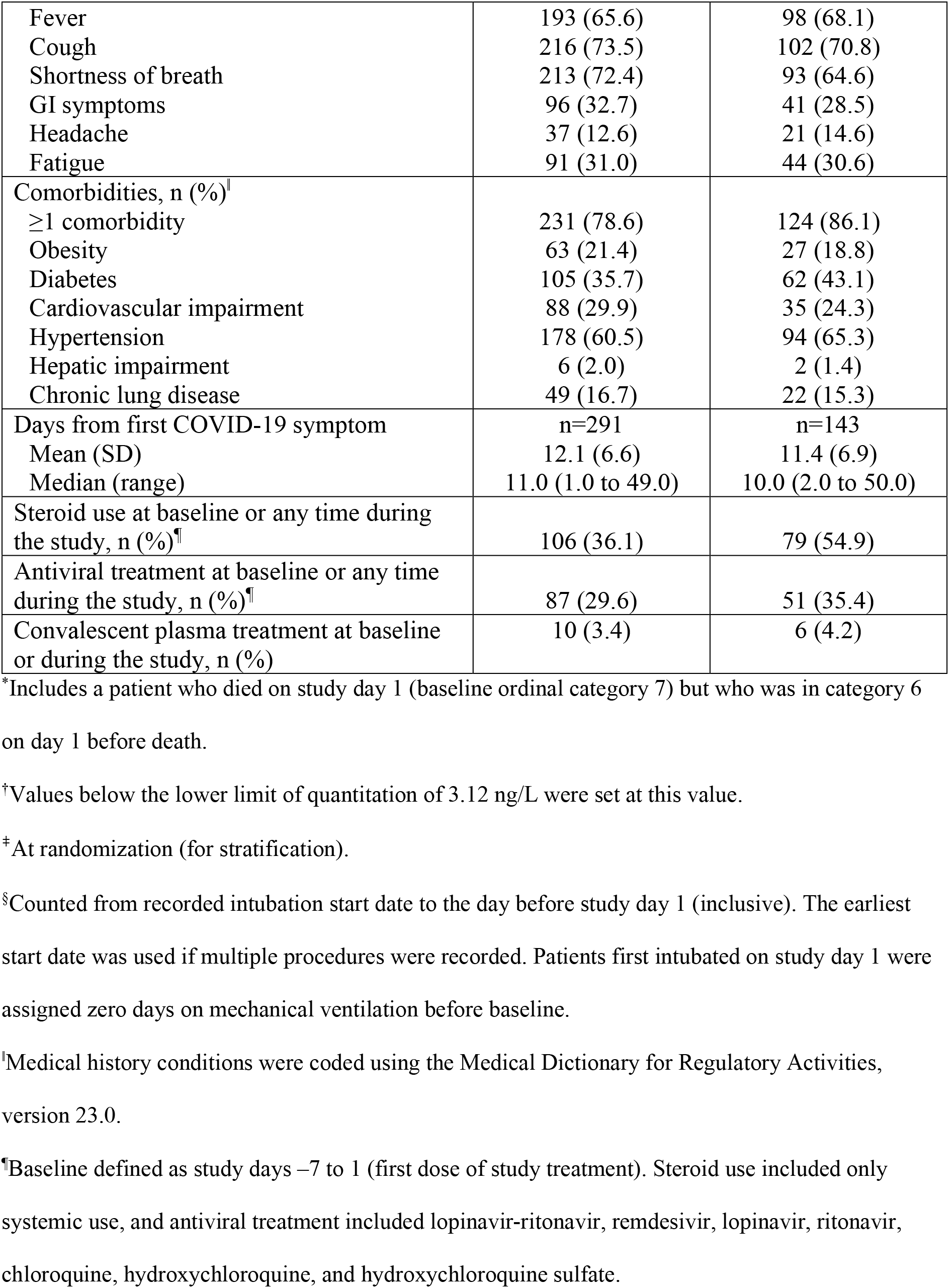
Baseline Demographics and Disease Characteristics

|  | **Tocilizumab**<br>**N=294** | **Placebo**<br>**N=144** |
| --- | --- | --- |
| Male, n (%) | 205 (69.7) | 101 (70.1) |
| Age, years, mean (SD) | 60.9 (14.6) | 60.6 (13.7) |
| Age category, years, n (%) |  |  |
| 18-64 | 163 (55.4) | 81 (56.3) |
| 65-84 | 117 (39.8) | 60 (41.7) |
| ≥85 | 14 (4.8) | 3 (2.1) |
| Weight, kg, mean (SD) | 88.9 (23.6) | n=143<br>88.1 (24.3) |
| Race, n (%) |  |  |
| American Indian or Alaska Native | 8 (2.7) | 5 (3.5) |
| Asian | 28 (9.5) | 10 (6.9) |
| Black or African American | 40 (13.6) | 26 (18.1) |
| Native Hawaiian or other Pacific Islander | 3 (1.0) | 5 (3.5) |
| White | 176 (59.9) | 76 (52.8) |
| Multiple | 0 | 1 (0.7) |
| Unknown | 39 (13.3) | 21 (14.6) |
| Region, n (%) |  |  |
| Europe | 120 (40.8) | 59 (41.0) |
| North America | 174 (59.2) | 85 (59.0) |
| NEWS2 score, mean (SD) | 7.1 (3.0) | 7.0 (3.0) |
| Ordinal scale for clinical status, n (%) |  |  |
| 2 | 9 (3.1) | 6 (4.2) |
| 3 | 78 (26.5) | 44 (30.6) |
| 4 | 94 (32.0) | 39 (27.1) |
| 5 | 45 (15.3) | 15 (10.4) |
| 6 | 68 (23.1) | 40 (27.8)* |
| Interleukin-6, ng/L<sup>†</sup> | n=233 | n=100 |
| Mean (SD) | 201.9 (418.4) | 195.4 (368.2) |
| Median (range) | 88.1 (3.1 to 4020) | 71.2 (3.1 to 2810) |
| CRP, mg/L | n=237 | n=125 |
| Mean (SD) | 168.4 (101.4) | 172.6 (114.0) |
| Median (range) | 157.2 (1.1 to 446.6) | 150.3 (1.6 to 499.6) |
| Ferritin, pmol/mL | n=241 | n=128 |
| Mean (SD) | 6891 (106736) | 4027 (45431) |
| Median (range) | 2.3 (0.0 to 1657000) | 2.2 (0.1 to 514000) |
| Mechanical ventilation, n (%)<sup>‡</sup> | 111 (37.8) | 54 (37.5) |
| Days on mechanical ventilation before baseline<sup>§</sup> among patients on mechanical ventilation at randomization | n=107 | n=51 |
| Mean (SD) | 5.1 (5.5) | 4.3 (4.5) |
| Median (range) | 3.0 (0.0 to 28.0) | 3.0 (0.0 to 20.0) |
| Symptoms at diagnosis, n (%) |  |  |
| Fever | 193 (65.6) | 98 (68.1) |
| Cough | 216 (73.5) | 102 (70.8) |
| Shortness of breath | 213 (72.4) | 93 (64.6) |
| GI symptoms | 96 (32.7) | 41 (28.5) |
| Headache | 37 (12.6) | 21 (14.6) |
| Fatigue | 91 (31.0) | 44 (30.6) |
| Comorbidities, n (%)<sup>†</sup> |  |  |
| ≥1 comorbidity | 231 (78.6) | 124 (86.1) |
| Obesity | 63 (21.4) | 27 (18.8) |
| Diabetes | 105 (35.7) | 62 (43.1) |
| Cardiovascular impairment | 88 (29.9) | 35 (24.3) |
| Hypertension | 178 (60.5) | 94 (65.3) |
| Hepatic impairment | 6 (2.0) | 2 (1.4) |
| Chronic lung disease | 49 (16.7) | 22 (15.3) |
| Days from first COVID-19 symptom | n=291 | n=143 |
| Mean (SD) | 12.1 (6.6) | 11.4 (6.9) |
| Median (range) | 11.0 (1.0 to 49.0) | 10.0 (2.0 to 50.0) |
| Steroid use at baseline or any time during the study, n (%)<sup>‡</sup> | 106 (36.1) | 79 (54.9) |
| Antiviral treatment at baseline or any time during the study, n (%)<sup>‡</sup> | 87 (29.6) | 51 (35.4) |
| Convalescent plasma treatment at baseline or during the study, n (%) | 10 (3.4) | 6 (4.2) |

\*Includes a patient who died on study day 1 (baseline ordinal category 7) but who was in category 6 on day 1 before death.
<sup>†</sup>Values below the lower limit of quantitation of 3.12 ng/L were set at this value.
<sup>‡</sup>At randomization (for stratification).
<sup>§</sup>Counted from recorded intubation start date to the day before study day 1 (inclusive). The earliest start date was used if multiple procedures were recorded. Patients first intubated on study day 1 were assigned zero days on mechanical ventilation before baseline.
<sup>‡</sup>Medical history conditions were coded using the Medical Dictionary for Regulatory Activities, version 23.0.
<sup>‡</sup>Baseline defined as study days –7 to 1 (first dose of study treatment). Steroid use included only systemic use, and antiviral treatment included lopinavir-ritonavir, remdesivir, lopinavir, ritonavir, chloroquine, hydroxychloroquine, and hydroxychloroquine sulfate.

### Primary Outcome

The primary endpoint was not met; clinical status on the 7-category ordinal scale at day 28 was not statistically significantly improved for tocilizumab versus placebo (van Elteren P=0.36). Median (95% CI) 7-category ordinal scale clinical status values at day 28 were 1.0 (1.0 to 1.0) for tocilizumab and 2.0 (1.0 to 4.0) for placebo; ordinal logistic regression odds ratio was 1.19 (0.81 to 1.76) (Table 2, Figure S1). Missing data were minimal for the primary endpoint of clinical status for the mITT population (3.7% tocilizumab, 2.1% placebo).

**Table 2.**
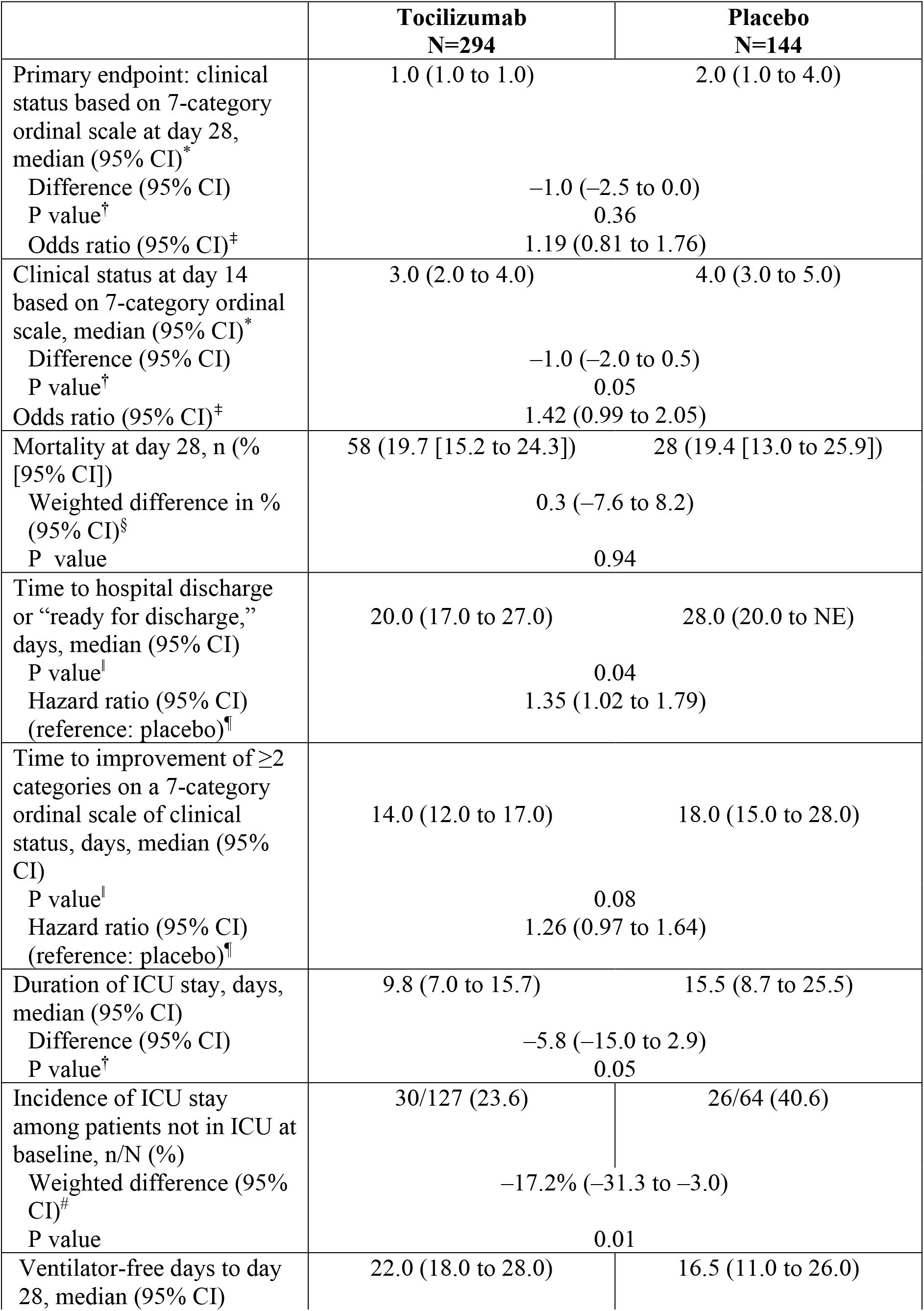
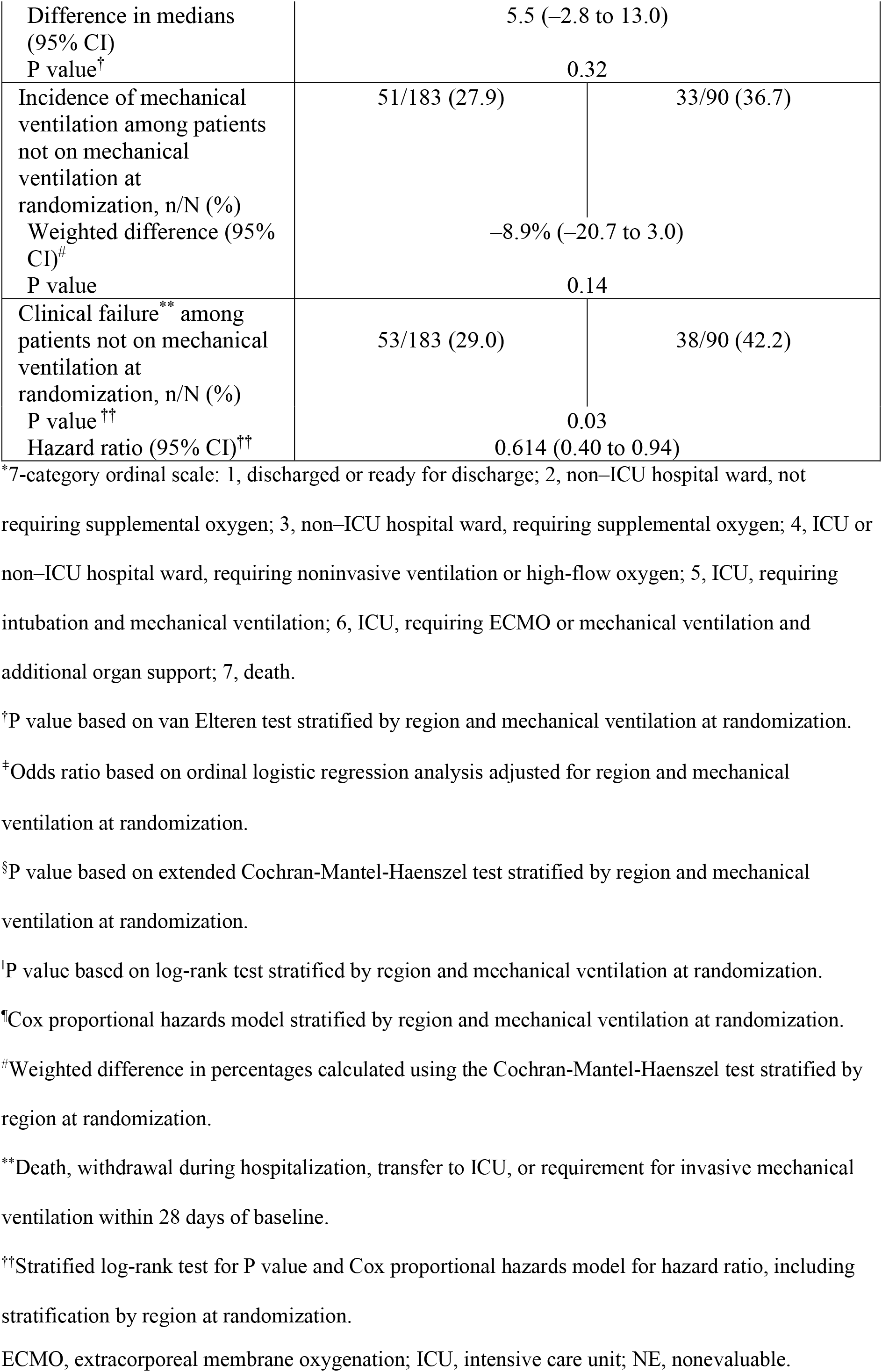
Efficacy Endpoints (modified-intention-to-treat population)

### Secondary Outcomes

All P values for secondary endpoints are nominal because the primary endpoint was not met. Median (95% CI) clinical status values on the 7-category ordinal scale at day 14 were 3.0 (2.0 to 4.0) in the tocilizumab arm and 4.0 (3.0 to 5.0) in the placebo arm (van Elteren P=0.05; odds ratio 1.42 [0.99 to 2.05]) (Table 2, Figure S2A). Fifty-eight patients (19.7%) in the tocilizumab arm and 28 (19.4%) in the placebo arm died by day 28 (weighted difference, 0.3% [95% CI –7.6% to 8.2%]; Cochran-Mantel-Haenszel P=0.94) (Table 2). The median (95% CI) number of ventilator-free days was 22.0 (18.0 to 28.0) with tocilizumab and 16.5 (11.0 to 26.0) with placebo (difference, 5.5 [–2.8 to 13.0]; van Elteren P=0.32) (Table 2). Median (95% CI) time to improvement from baseline in ≥2 categories on the 7-category ordinal scale was 14 days (12 to 17) in the tocilizumab arm and 18 days (15 to 28) in the placebo arm (log rank P=0.08; Cox proportional hazards ratio 1.26 [95% CI 0.97 to 1.64]) (Table 2, Figure 2A). Median (95% CI) time to hospital discharge/ready for discharge was 20 days (17 to 27) in the tocilizumab arm and 28 days (20 to nonevaluable) in the placebo arm (log rank P=0.04; Cox proportional hazards ratio 1.35 [1.02 to 1.79]) (Table 2, Figure 2B). Median duration of ICU stay was 9.8 days in the tocilizumab arm and 15.5 days in the placebo arm (difference, –5.8 days [95% CI −15.0 to 2.9]; van Elteren P=0.05) (Table 2). Cumulative incidences of time to improvement in clinical status, time to hospital discharge/ready for discharge, and mortality are shown in Figure S3.

**Figure 2.**
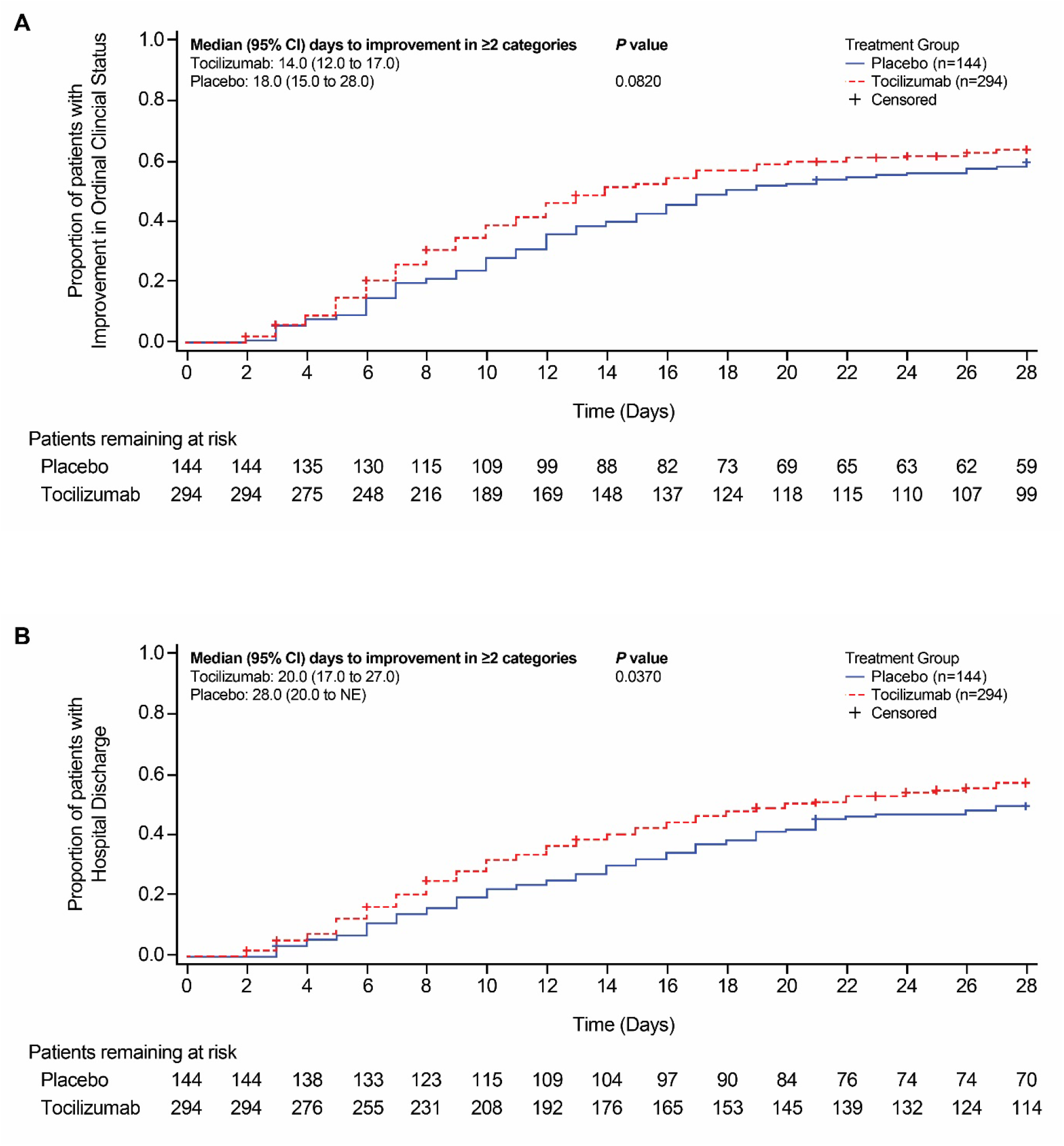
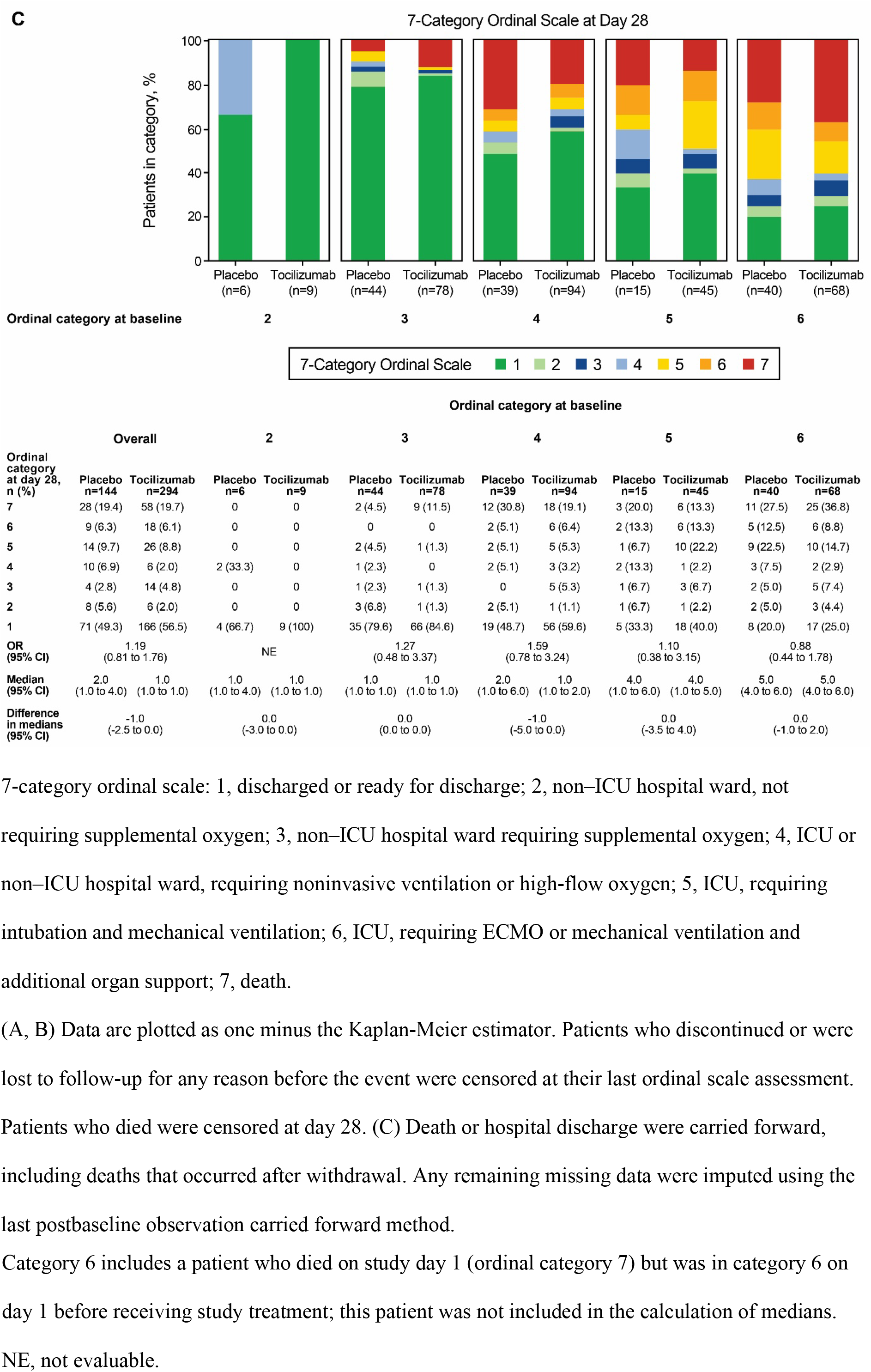
(A) time to improvement of ≥2 ordinal scale categories relative to baseline, (B) time to hospital discharge/ready for discharge to day 28, and (C) clinical status assessed using 7-category ordinal scale at day 28 according to baseline ordinal scale category (modified-intention-to-treat population for all analyses).

### Subgroup Analyses

Among 122 patients with baseline ordinal scale category 3 and 133 patients with baseline category 4, odds ratios (95% CIs) for improved clinical status at day 28 were 1.27 (0.48 to 3.37) and 1.59 (0.78 to 3.24), respectively; among 60 patients with baseline category 5 and 108 patients with baseline category 6, odds ratios for improved clinical status were 1.10 (0.38 to 3.15) and 0.88 (0.44 to 1.78), respectively (Figure 2C, Figure S2B). There was no significant difference in clinical status on the ordinal scale at day 28 between tocilizumab and placebo among patients mechanically ventilated at randomization (median [95% CI], 5.0 [3.0 to 5.0] (n=111) vs 5.0 [4.0 to 6.0] (n=54); odds ratio 1.04 [0.58 to 1.85]) or those not mechanically ventilated at randomization (1.0 [1.0 to 1.0] (n=183) vs 1.0 [1.0 to 1.0] (n=90); odds ratio 1.34 [0.79 to 2.27]) (Figure S4).

The incidence of mechanical ventilation among patients not mechanically ventilated at randomization was 27.9% (51/183) in the tocilizumab arm and 36.7% (33/90) in the placebo arm (weighted difference, –8.9% [95% CI −20.7% to 3.0%]; Cochran-Mantel-Haenszel nominal P=0.14). The incidence of ICU transfer among patients not in ICUs at baseline was 23.6% (30/127) in the tocilizumab arm and 40.6% (26/64) in the placebo arm (weighted difference, –17.2% [95% CI −31.3% to −3.0%]; Cochran-Mantel-Haenszel nominal P=0.01). In post hoc analysis, among patients not mechanically ventilated at randomization, 53 of 183 (29.0%) in the tocilizumab arm and 38 of 90 (42.2%) in the placebo arm experienced clinical failure (includes those who died, withdrew during hospitalization, were transferred to an ICU, or required invasive mechanical ventilation within 28 days of baseline, as defined in Methods) (hazard ratio 0.614; 95% CI 0.40 to 0.94; nominal P=0.03).

### Safety

In the safety population, adverse events were reported in 77.3% of 295 patients in the tocilizumab arm and 81.1% of 143 patients in the placebo arm through day 28 (Table 3); serious adverse events were reported in 34.9% and 38.5%, respectively. Fatal events occurred in 58 patients (19.7%) in the tocilizumab arm and 28 (19.6%) in the placebo arm through day 28. The most commonly reported cause of death was COVID-19 pneumonia. Adverse events of special interest for tocilizumab were generally balanced between treatment arms. No tocilizumab-treated patients experienced anaphylaxis. Seventy-six serious infections were reported in 62 patients (21.0%) in the tocilizumab arm and 49 in 37 patients (25.9%) in the placebo arm through day 28. Similar proportions of patients in each treatment arm experienced adverse events and serious adverse events through the clinical cutoff date of June 24, 2020 (Table S1).

**Table 3.**
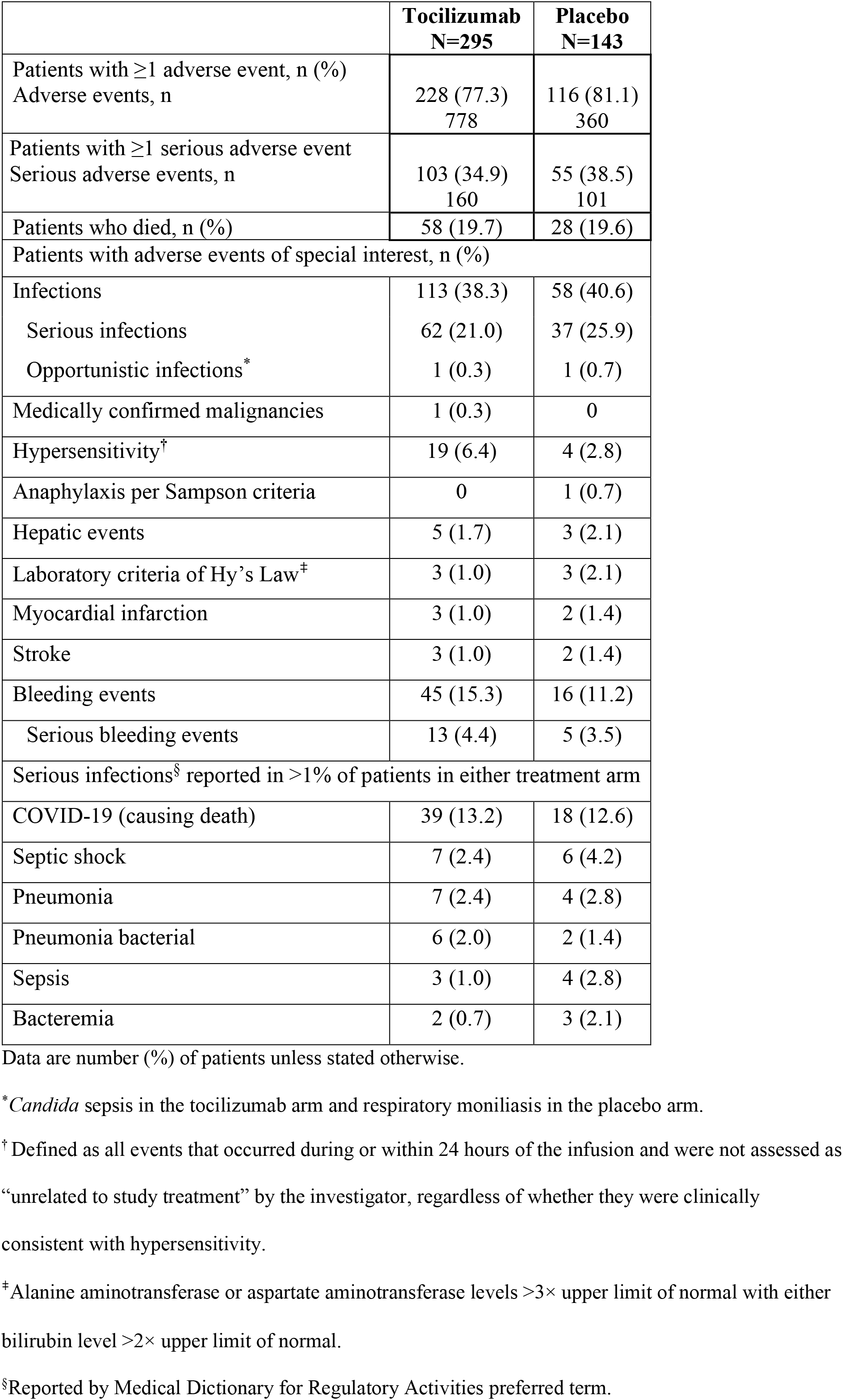
Safety to day 28 (safety population)

## DISCUSSION

COVACTA, the first randomized, double-blind, placebo-controlled trial of tocilizumab in COVID-19 pneumonia, included patients from 9 countries. The primary endpoint was not met; there was no significant difference between tocilizumab plus standard care and placebo plus standard care in clinical status assessed using a 7-category ordinal scale at day 28, and no mortality benefit was demonstrated. However, tocilizumab appeared to be safe, and potentially clinically meaningful benefits were identified in time to hospital discharge/ready for discharge and duration of ICU stay. Among patients not mechanically ventilated at randomization, fewer treatment failures (progression to mechanical ventilation, ICU admission, or death) occurred in tocilizumab-treated than placebo-treated patients. Because the primary endpoint of the study was not met, these findings require validation in additional studies. Adverse events, including those of special interest for tocilizumab (bleeding events, hepatic events, cardiac events), were generally balanced between tocilizumab and placebo, and incidences of infections or serious infections were lower in the tocilizumab arm.

The design and conduct of clinical trials in patients with COVID-19 present unique challenges and limitations. The COVACTA study population was intentionally chosen to be heterogeneous with regard to patient characteristics, previous/concurrent treatments, and disease severity to allow assessment of potential benefit across a broad range of patients and to reflect real-world practice in the expanding pandemic. Despite this heterogeneity, the proportion of patients discharged or ready for discharge by day 28 was higher in the tocilizumab arm than the placebo arm across the baseline ordinal scale of clinical status categories, whereas no consistent pattern was observed for mortality. The lack of standardized treatment across study sites and countries is an important limitation of this study considering potential interactions with antivirals and steroids. More patients in the placebo arm than the tocilizumab arm received concomitant steroids, which might have created bias toward lower mortality in the placebo arm^22^; however, this imbalance is unlikely to have obscured a significant treatment effect because the mortality rate was similar between treatment arms regardless of steroid use and was higher in patients who received steroids in both treatment arms than in those who did not (Table S2). Since our study was initiated, standard care treatment and understanding of the natural history of COVID-19 and its associated complications have evolved substantially. Based on current knowledge, optimal endpoints for clinical trials and effective treatments are likely to be different for different stages of disease. Future trials should be more narrowly focused or much larger to allow for further stratification based on disease severity and other baseline characteristics.

Results of this study must be interpreted in the context of therapies for severe COVID-19. Among treatments for patients hospitalized with COVID-19 investigated in randomized controlled trials, dexamethasone reduced mortality in patients receiving mechanical ventilation or supplemental oxygen at randomization, but not in patients not receiving respiratory support.^22^ Remdesivir shortened time to recovery, but there was no statistically significant difference in 14-day mortality.^23^ Clinical trials investigating potential treatments—including other antivirals, anti-inflammatories, other targeted immunomodulators (sarilumab, anakinra, baricitinib, canakinumab), anticoagulants, and antifibrotics (tyrosine kinase inhibitors)—are underway,^24^ but the urgent need for effective treatments remains. In the absence of a more effective therapy, treatments such as tocilizumab, which this study suggests might hasten recovery and decrease the need for intensive care without increasing the risk for infections, serious infections, or other adverse events, might be clinically useful, even without a demonstrated mortality benefit.

Additional studies are ongoing and might expand the findings of COVACTA and address outstanding scientifically and medically relevant questions regarding the risk/benefit profile of tocilizumab in COVID-19 in more narrowly defined patient populations and in conjunction with current treatments.

## Data Availability

Qualified researchers may request access to individual patient level data through the clinical study data request platform (www.clinicalstudydatarequest.com). Further details on Roche's criteria for eligible studies are available here (https://clinicalstudydatarequest.com/Study-Sponsors/Study-Sponsors-Roche.aspx). For further details on Roche's Global Policy on the Sharing of Clinical Information and how to request access to related clinical study documents, see here (https://www.roche.com/research_and_development/who_we_are_how_we_work/clinical_trials/our_commitment_to_data_sharing.htm)

https://www.roche.com/research_and_development/who_we_are_how_we_work/clinical_trials/our_commitment_to_data_sharing.htm

## Acknowledgments

The first draft of the manuscript was prepared by Larry Tsai, M.D., with writing support provided by Sara Duggan, Ph.D., of ApotheCom, funded by F. Hoffmann-La Roche Ltd. The data were analyzed by Helen Spotswood, Ph.D., Sophie Dimonaco, M.Sc., and Emily Graham, Ph.D., funded by Roche. The study was funded by F. Hoffmann-La Roche Ltd and funded in part with federal funds received from the Department of Health and Human Services, Office of the Assistant Secretary for Preparedness and Response, Biomedical Advanced Research and Development Authority, under OT number HHSO100201800036C.

## Conflicts of Interest

I.O.R. received a grant from Roche/Genentech during the conduct of the study; a grant and personal fees from Genentech outside the submitted work; and personal fees from Boehringer and Bristol-Myers Squibb outside the submitted work.

N.B.’s institution received grant support from Roche/Genentech during the conduct of the study.

M.W., M.S.A., N.C., I.S.D., S.S., T.Y., A.C.G., and D.J.D.L.Z. have nothing to disclose.

R.G. has received consulting fees from F. Hoffmann-La Roche outside the submitted work.

B.D.H. has received speaker bureau fees from Kite Pharmaceuticals outside the submitted work.

S.B. has received personal fees from Gilead Sciences, Roche, and ViiV Healthcare outside the submitted work.

D.S.’s institution received grant support from Roche/Genentech during the conduct of the study.

L.D.S. received nonfinancial support from F. Hoffmann-La Roche Ltd during the conduct of the study.

A.U.’s institution received grant support from Roche/Genentech during the conduct of the study, and he has received personal fees from Gilead Sciences outside the submitted work.

M.B. and L.T. received a grant from Biomedical Advanced Research and Development Authority for the COVACTA study; are employees of Roche/Genentech; and have filed a patent for a method of treating pneumonia, including COVID-19 pneumonia, with an IL-6 antagonist.

S.D. received a grant from Biomedical Advanced Research and Development Authority for the COVACTA study and is an employee and a shareholder of Roche Products Ltd.

E.G. and B.M. are employees of Roche Products Ltd.

H.S. is an employee and a shareholder of Roche Products Ltd.

A.M.’s institution received grant support from Roche/Genentech during the conduct of the study; he has received funding from the National Institutes of Health outside the submitted work and medical education from Merck and Livanova outside the submitted work.

## Notes

### Clinical Trial

NCT04320615

### Author Declarations

Informed consent was obtained for all enrolled patients. The study was conducted in accordance with the International Council for Harmonization E6 guideline for good clinical practice and the Declaration of Helsinki or local regulations, whichever afforded greater patient protection. The protocol was reviewed by institutional review boards or ethics committees.

